# The magnitude, determinants, and outcome of shock among pediatric patients: A cross-sectional hospital-based study

**DOI:** 10.1101/2023.10.27.23297683

**Authors:** Mebrahtu G. Kidanu, Engdaeshet Tazebe, Alemseged Birhane, Marta Yemane, Mebrihit M. Kahsay, Mebrahtu G. Tedla

## Abstract

**Background:** Pediatric shock, a potentially fatal illness, develops after a systemic circulatory system failure. It appears to be a common emergency in children and produces substantial morbidity and mortality particularly if there is no early identification and therapy. The extent and causes of shock-induced death among children in Ethiopia have not been sufficiently studied.

**Objective:** This study was conducted to evaluate the severity, determinants, and prognosis of shock in pediatric patients who visited Ayder Comprehensive Specialized Hospital in Tigray, Northern Ethiopia.

**Methods:** From October 1st, 2020, to July 30th, 2022, an observational cross-sectional study was carried out at Ayder Comprehensive Specialized Hospital. The study included 132 children from the age of 1 month to 18 years. According to pediatric advanced life support guidelines, shock was diagnosed among patients. To gather information, a pretested questionnaire was employed. To examine the relationship between the independent variables and shock outcome, bivariate logistic regression was performed, and statistical significance was defined as a P-value of 0.05 or lower.

**Result:** The prevalence of shock was 2.2%. This study revealed 70.4 % decompensated stage of shock. Mortality rate of shock was 45.5% (95% CI: 37.1-53.8). A delayed presentation by more than one week with an adjusted odd ratio(AOR) of 16.9 (95% CI:2.3-123), type of shock other than hypovolemic shock with AOR of 8.3 (95% CI: 1.4-48), stage of shock with AOR of 27.8 (95% CI: 2.8-157), requirement of mechanical ventilation with AOR of 11 (95% CI:2.6-53) and length of hospital stay less than three days with AOR of 9 (95% CI: 1.7-48) were identified as a predictor of mortality by shock in children.

**Conclusion:** According to this study, shock causes a higher rate of child mortality. Independent predictors of mortality included delayed presentation, shock type, shock stage, need for mechanical ventilation, and brief hospital stay (less than three days).

## Background

The body’s incapacity to supply enough oxygen to meet the metabolic needs of essential organs and tissues causes an acute state of shock. Adverse vascular, inflammatory, metabolic, cellular, endocrine, and systemic responses exacerbate physiologic instability if insufficient tissue perfusion continues [1]. Hypovolemic, distributive, cardiogenic, obstructive, and dissociative types are five broad classifications of shock [1,2]. Intensive treatment must be started quickly to manage shock in children. Most pediatric hospital deaths frequently happen within the first 24 hours of admission. Early goal-directed therapy for shock aimed at improving vital organ function and physiologic indications of perfusion within the first six hours of treatment, combined with an aggressive, systematic strategy to resuscitation [3]. Multiple organ system failure is the primary complication of shock and is linked to a higher death rate and longer hospital stays in survivors [1, 2].

In developed countries, shock is thought to affect 2% of all hospitalized patients, and the death rate varies according to the type of shock and clinical circumstances [1]. However, shock in children is associated with a high morbidity and mortality rate in low– and middle-income countries, with prevalence ranging from 1.5 to 44.3% and mortality from 3.9 to 33.3%. Most deaths happen during the first 24 hours of admission, and most of them are avoidable [4].

Preliminary studies and reports from hospitals in Ethiopia showed that there is high prevalence of shock and is associated with high mortality. For example, a cross-sectional study on admitted and outcome patients of children at a hospital located in South-Western Ethiopia showed shock in children was the leading medical cases which accounts for 17% of the admissions and a high mortality rate of up to 50% [5]. There is paucity of data on short term outcome of shock in children in Ethiopia, in particular in our study area. Therefore, this study was conducted to assess the magnitude of pediatric shock and to identify the factors associated with mortality among children visited to pediatric emergency outpatient unit of the hospital.

## 2 Methods and Materials

### Study area

Ayder Comprehensive Specialized Hospital is a referral and teaching hospital found in Ethiopia’s Tigray region, Mekelle and this study was conducted from October 1, 2020, to July 30, 2022.

### Study design

Institution based cross-section study was conducted and patients aged between 1 month to 18 years and who have visited to the hospital, department of pediatrics and child health.

### Inclusion and exclusion criteria

Children from the age of 1 month up to 18 years old teenagers were included in the study. Patients or guardians who received medical advice were included in the study. Whereas those who didn’t receive medical advice were excluded from the study.

### Sample size determination and sampling procedure

The sample size was determined using a single population proportion formula with a 95% confidence level, 5% margin of error as follows: n= <u>(Z)² × P (1 p)</u> / (d)² = (1.96)² × 0.09(1-0.09) ÷(0.05)² = 126; where: **n**: is the minimum sample size required**; p**: prevalence of shock in children **; Z**: is the standard normal variable at (1-α) % confidence level and α is mostly 0.05 i.e. with 95% CI (z=1.96)**; d**: is the margin of error to be tolerated (5%); n = 139(10%).

### Data collection tools and procedures

A semi-structured questionnaire that had been pre-tested was used to gather data. After being translated into Tigrigna, the local language, the questionnaire was pretested in English. Based on the PALS criteria, information was gathered from every child who had been diagnosed with shock. Age-specific ranges were used to interpret investigations and vital signs, and the WHO curve was also used to interpret anthropometry results. The use of pretested questioners helped to ensure the accuracy of the data. Ten patients who were enrolled in the study were given a pretest version of the questionnaire, and the language’s clarity was examined. Each questionnaire was examined by the lead investigator prior to data entry.

### Data processing and analysis

Data were first verified and coded before being put into Epi Data Manager version 4.6.0.2. From there, they were exported to the Statistical Package and Service Solution (SPSS) version 23 for analysis. Frequency and percentages were used to express category variables. After being verified to have a normal distribution, those continuous variables were expressed as the mean plus standard deviation and median. First, bivariate logistic regression analysis was used to evaluate the factors that influence the short-term outcome of shock in children.

### Ethical approval and consent to participate

The Institutional Review Board (IRB) of Mekelle University’s College of Health Sciences granted ethical approval (MU-IRB 1998/2022: ethical approval number). Consent was obtained from all guardians of study participants, both verbally and in writing. All guardians of study participants received information on the purpose of the study, its contents, and their interest to decline or stop participating in the study at any time.

## 3. Result

Out of a total 6112 visits to the Pediatric Emergency Outpatient Department (PEOPD), 134 participants were diagnosed with shock, and this gives a point prevalence 2.2%. The Socio-demographic characteristics of study participants and caregivers were also analyzed, and the median age of the study participants was 6.45 years. Thirty-seven (28%) were infants, 38(28.8%) were between the age of 1 and 5 years and 57(43.2%) were above 5 years old. The male to female ratio was 1.3: 1. More than half (55.3%) of the participants were from urban area, 30 (22.7%) of attendants were not educated and 88 (66.7%) of the participants were referred from other facility (Table 1).

**Table 1:** Socio-demographic characteristics of study participants and caregivers among pediatric shock patients.

| Variables | Category | Count(n) | Percentage (%) |
| --- | --- | --- | --- |
| **Sex of the children** | Male | 75 | 56.8% |
|  | Female | 57 | 43.2% |
| **Residence** | Urban | 73 | 55.3% |
|  | Rural | 59 | 44.7% |
| **Age of participant** | 1month-11 month | 37 | 28% |
|  | 1-5 years | 38 | 28.8% |
|  | Above 5 years | 57 | 43.2% |
| **Mothers' educational status** | No formal education | 30 | 22.7% |
|  | Primary | 52 | 39.3% |
|  | Secondary | 36 | 27.4% |
|  | Tertiary | 14 | 10.6% |
| **Referred from other health facility** | Yes | 88 | 66.7% |
|  | No | 44 | 33.3% |

### 3.1. Clinical profile and laboratory result of participants

Among the patients included in the study, the most common symptom was vomiting;103(78%), followed by altered mentation;91 (68.9%), fast breathing; 88(66.7%), fever; 80(60.6%), diarrhea; 76(57.6%). A total of 72(54.4%) of them were presented after one week of illness, 34(25.8%) were presented within two days of illness. From the total of 132 cases, one third (31.8%) of them had chronic medical illness. Those include cardiac 12(28.6%), hematologic disease 11(26.2%), diabetes mellitus 9(21.4%), epilepsy 7(16.7%), respiratory viral infection 2(4.8%) and 1(2.3%) with brain tumor. The most common clinical sign noticed was tachycardia 115(87.1%), feeble pulse volume 111(84.1%), positive sign of dehydration 98(74.2%), tachypnea 94(71.2%), saturation below 90% in 88(66.7%), decrease urine output in 84 (63.4%), pallor in 68(51.5%). Blood pressure measurement was taken for 58(43.9%) and it was unrecordable in more than 20(35.1%) of them. Nutritional assessment showed that severe wasting was noticed in 55(41.7%) of the total. Mean Glasgow coma scale (GCS) of participants was 12.5 (SD±2.6) and nineteen (14.4%) patients were comatose at presentation (Table 2).

**Table 2:**
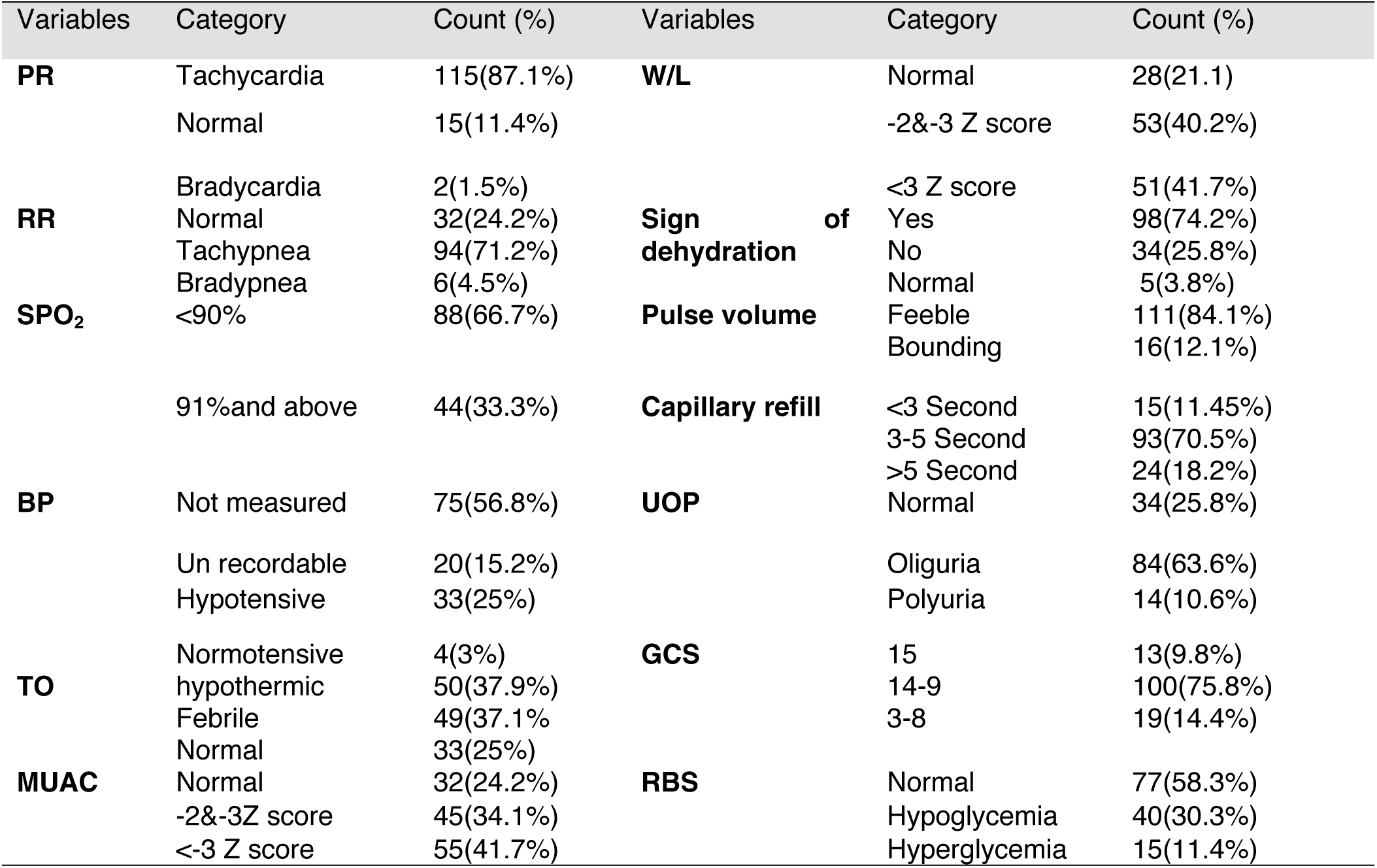
Physical finding and measurements of pediatric shock patients.

A complete blood count was done for 87(65.9%) of the cases. Hemoglobin blood test was performed in 93(70.5%) cases and the result showed 57(61.3%) of them had low hemoglobin level. Creatinine level test in 94(71.2%) cases showed an elevation in more than half 54(57.4%) of them. Liver function test (LFT) was performed for only one-third participants, and it was elevated in nearly half of them. Serum electrolyte (sodium and potassium) test was also performed in less than two third (64.4%) of participants but only eleven participants had serum calcium determination and nine of them showed hypocalcemia (Table 3). Electrolyte abnormality and stage of shock pediatric patient was also assessed (Table 4).

**Table 3:**
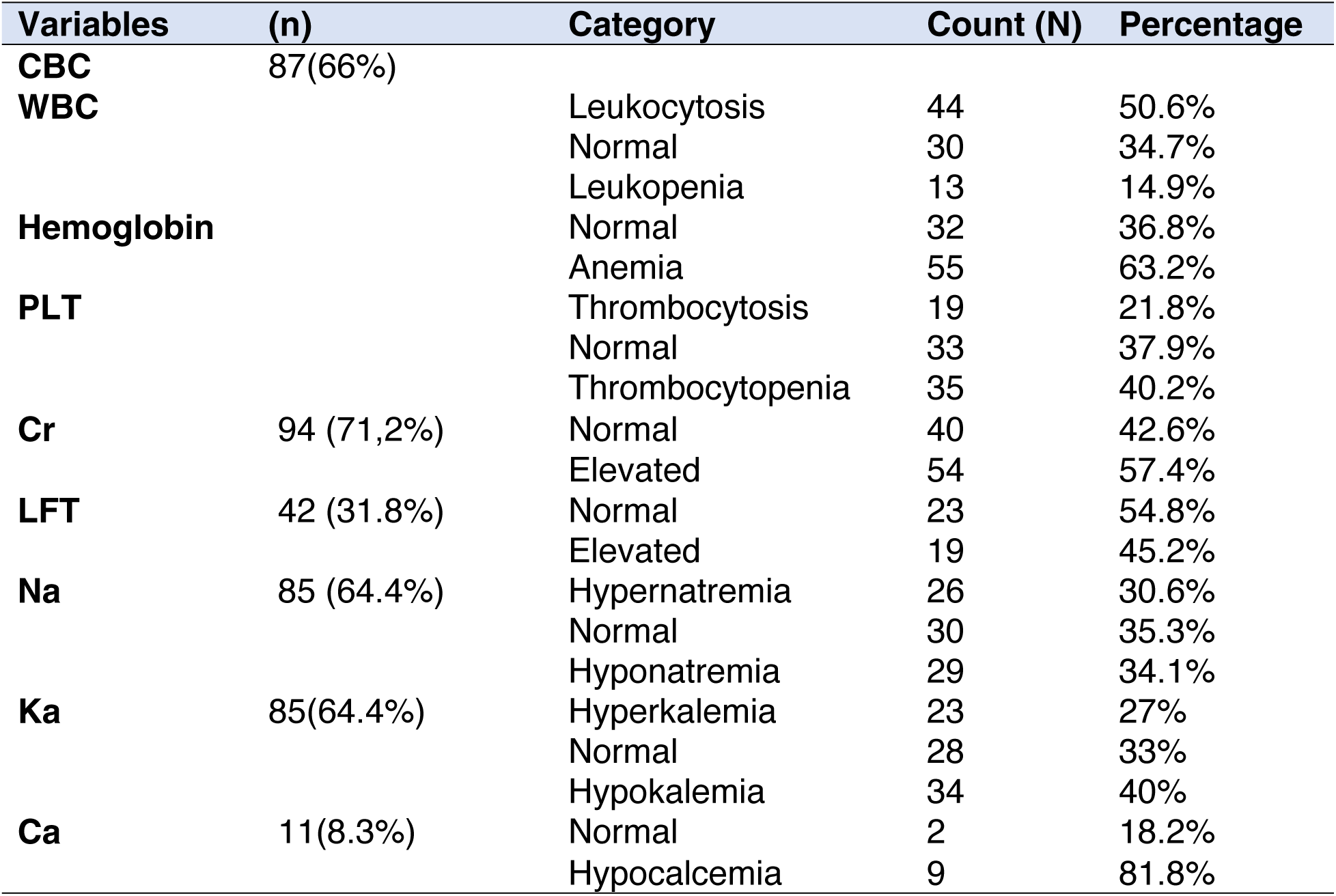
Laboratory parameters of pediatric shock patients.

**Table 4:** Electrolyte abnormality and stage of shock of pediatric patient.

| Variables | Compensated |  | Decompensated |  |
| --- | --- | --- | --- | --- |
|  | Count(n) | % | Count(n) | % |
| Hypernatremia | 6 | 23.1 | 20 | 76.9 |
| Hyponatremia | 13 | 44.8 | 16 | 55.2 |
| hyperkalemia | 8 | 34.8 | 15 | 65.2 |
| Hypokalemia | 13 | 38.2 | 21 | 61.8 |
| Hypocalcemia | 3 | 33.3 | 6 | 66.7 |

### 3.2. Type, Stage and Management of shock

The most common type of shock was hypovolemic shock (53%), followed by septic shock (34.4%), mixed (6.8%), cardiogenic and anaphylactic shock each account for 2.3% respectively. Only one patient presented with obstructive shock. Out of the total 132 cases,93(70.5%) of them were in decompensated stage. 44(33.3%) of participants had multiorgan failure. During the management of shock, 76(57.6%) patients received crystalloid boluses and 56(42.4%) cases were given both crystalloid and colloids. Out of the 60(45.5%) cases who required inotropic, 61.7% of them took both dopamine and adrenalin, 18(30%) dopamine only and 5(8.3%) adrenalin only, 37.9% of participants received steroid. 35.6% of the cases required mechanical ventilation. The time gap for the initiation of fluid after the patient arrived was determined and the mean duration was 24±10 minutes. The frequency of re-evaluation was every median of 1hr. The median length of hospital stay was 7 (Table 5 and Figure 2).

**Figure 1:**
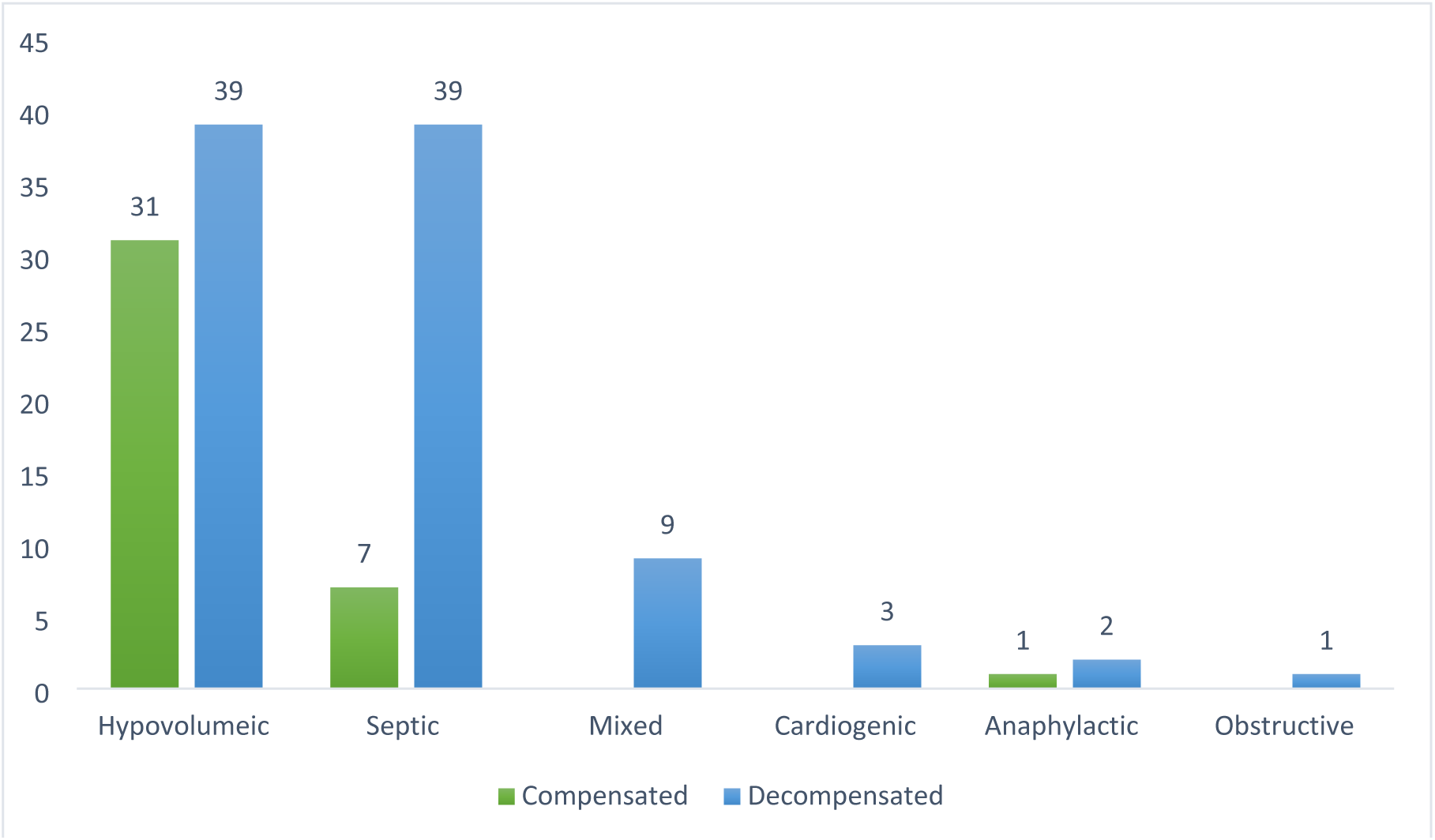
Type and stage of shock children.

**Figure 2:**
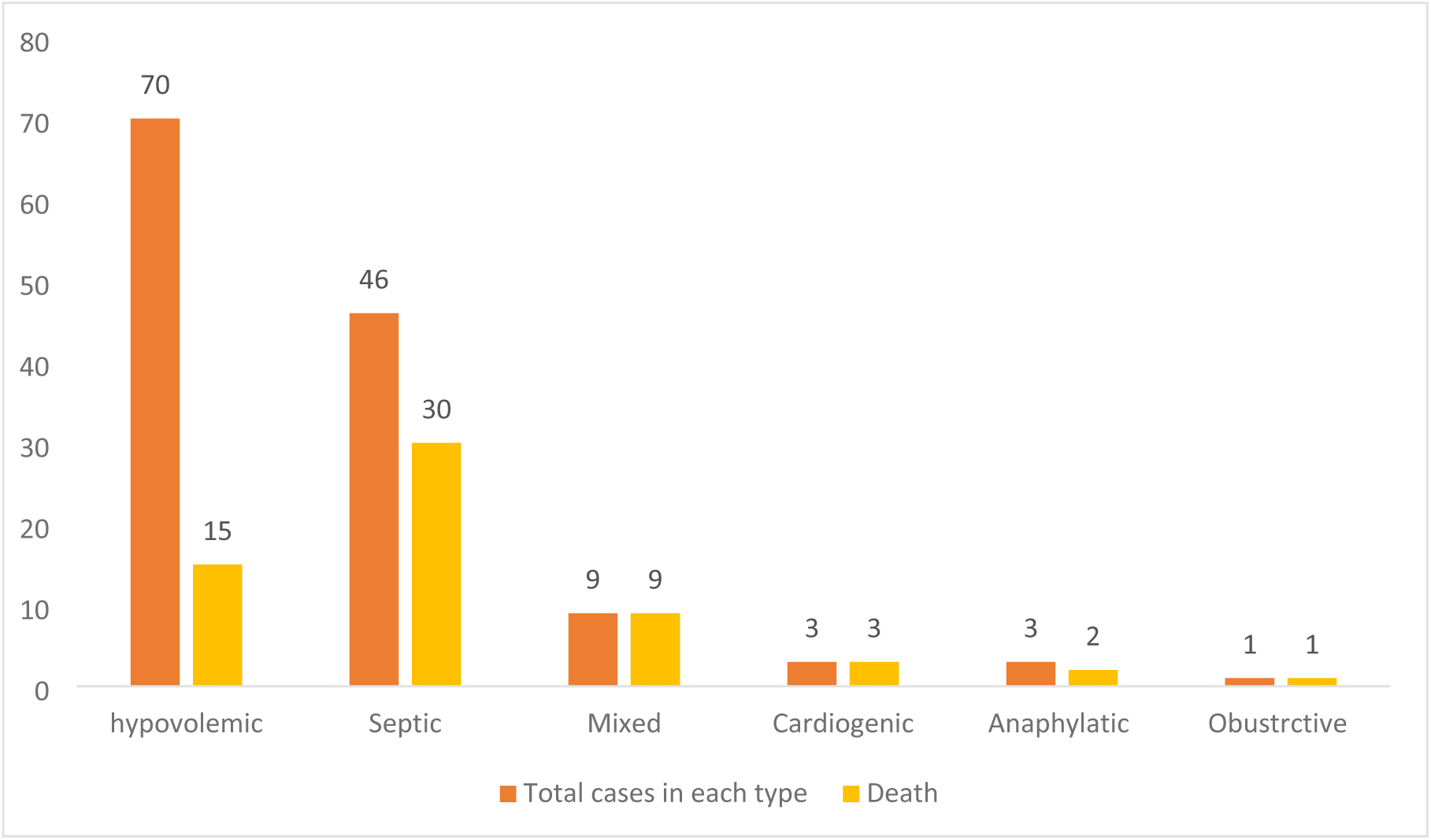
Type versus outcome of pediatric shock patients.

**Table 5:** Type, stage and management of pediatric shock patients.

| Variables | Characteristics | Count(n) | Percentage |
| --- | --- | --- | --- |
| Type of shock | Hypovolemic | 70 | 53% |
|  | Septic | 46 | 34.8% |
|  | Mixed | 9 | 6.8% |
|  | Cardiogenic | 3 | 2.3% |
|  | Anaphylactic | 3 | 2.3% |
|  | Obstructive | 1 | 0.8% |
| Stage of shock | Compensated | 39 | 29.5% |
|  | Decompensated | 93 | 70.5% |
| Multi organ dysfunction syndrome | Yes | 44 | 33.3% |
|  | No | 88 | 66.7% |
| Fluids administration | Crystalloids only | 76 | 57.6% |
|  | Crystalloids and colloid | 56 | 42.4% |
| Antibiotic administration | Started just on arrival | 45 | 35.2% |
|  | He/she was on antibiotic | 83 | 64.8% |
| Inotropic administration | Yes | 60 | 45.5% |
|  | No | 72 | 54.5% |
| Steroid administration | Yes | 50 | 37.9% |
|  | No | 82 | 62.1% |
| Invasive ventilation provided | Yes | 47 | 35.6% |
|  | No | 85 | 64.4% |
| Patient outcome | Improved | 72 | 54.5% |
|  | Dead | 60 | 45.5 % |

### 3.3. Outcome of shock in pediatric patient

The overall mortality of shock in this study was 45.5% (95% CI, 37.1-53.8). Out of those, 64.5% were in decompensated stage of shock. The proportion of death was higher in those above five years old (43.3%) followed by infants (30%) and 26.7% in 1-5 years old. But the case fatality rate was higher among those infants (48.6%), followed in those above five years 45.6% and 42.1% in 1-5 years. 70% of death were referred from other health facility. Mortality of mixed type cardiogenic and obstructive type of shock were 100%. Case fatality rate of septic shock was 65.2% and 21.4% for hypovolemic shock. 50% of mortality was due to septic shock (Figure 2).

### 3.4. Predictors of mortality shock among pediatric patients

A binary logistic regression analysis was performed to identify independent predictor or causes of mortality by shock among pediatric patients(Table 6) and our result revealed delayed presentation by more than one week with an adjusted odd ratio(AOR) of 16.9 (95% CI:2.3-123), type of shock other than hypovolemic shock with AOR of 8.3 (95% CI: 1.4-48), stage of shock with AOR of 27.8 (95% CI: 2.8-157), requirement of mechanical ventilation with AOR of 11 (95% CI:2.6-53) and length of hospital stay less than three days with AOR of 9 (95% CI: 1.7-48).

**Table 6:**
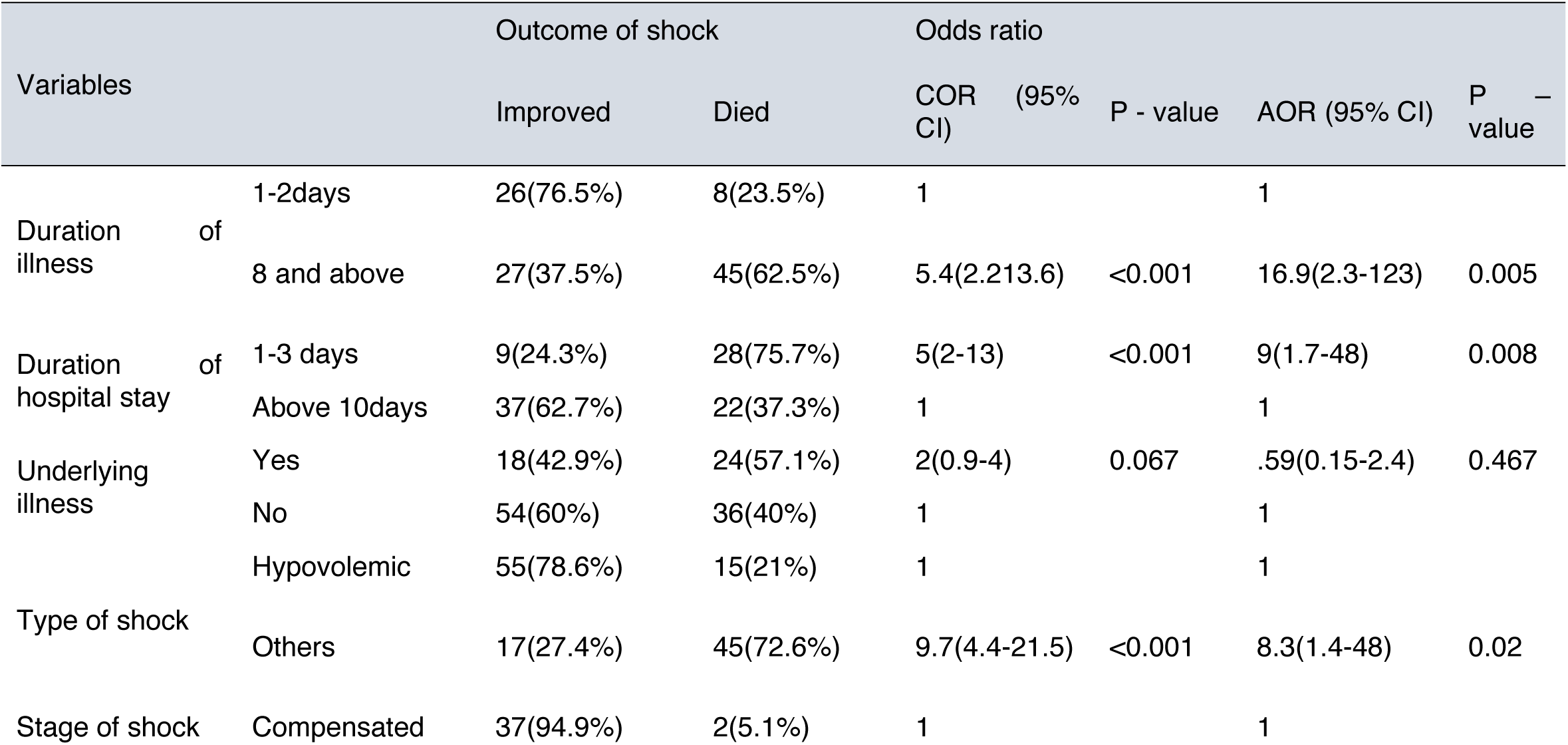
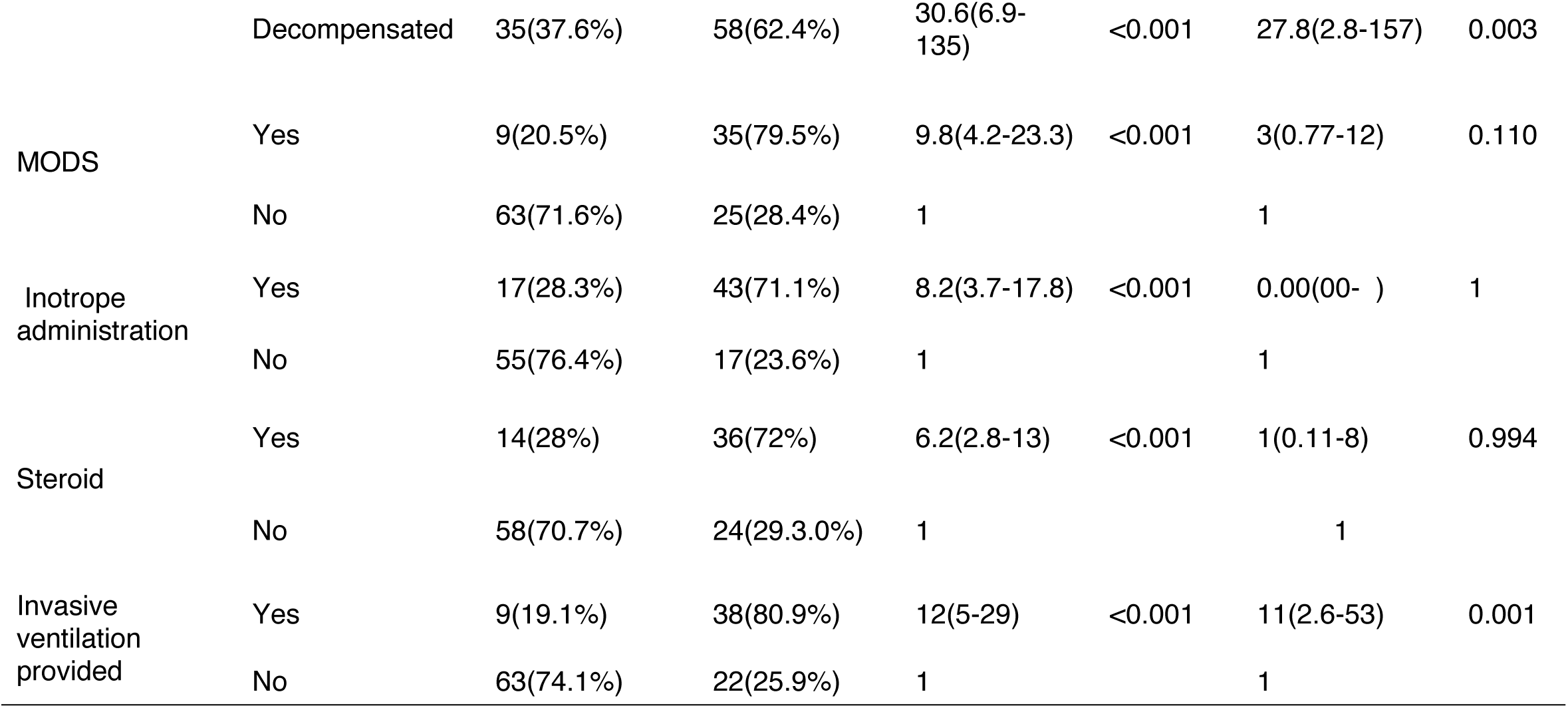
Assessing predictors of mortality among pediatric shock patient using binary logistic regression.

## Discussion

In this study, we have assessed and confirmed the magnitude and factors associated with mortality of shock in children who visited Ayder Comprehensive Specialized Hospital. The prevalence of shock was 2.2%. S similar studies on the prevalence of shock among all hospitalized infants, children and adult has been reported in developed countries which was 2% [1]. However, this prevalence is lower comparing to reports from another countries; 4.3% [6]; 9% [7], and 2.8% [8]. In this study, the most common type of shock was identified as hypovolemic shock (53%). Only one patient with obstructive shock with massive pericardial and plural fluid was found. Our finding is supported by previous studies conducted elsewhere who have shown the most common type of shock was hypovolemic shock that accounted for 45.9% cases followed by septic type (34.6%), cardiogenic type (17.3%) and distributed type (2%) [8]. However, previous studies showed prevalence based on type of shocks as septic 28%, distributive 22%, septic(cardiogenic) type 17.5%, hypovolemic15.7%, and cardiogenic shock 12% [7]. In our study there were 93(70.5 %) with decompensated stage of shock, which is higher than previous studies who 40 [8], and 57.9 % [7].

In this study those with mixed, cardiogenic and obstructive type of shock were all in decompensated stage of shock. The decompensation in hypovolemic and septic were 55.7% (39/70) and 84.7 % (39/46) respectively. This could be because of the fact that 66.7% (88) of children included in the study were referrals from another hospital, with associated delay in recognition or transfer to our hospital for better management. Of the 42(31.8%) children who received liver function test, 19(45.2%) have elevated values, and 13(68.4%) of them were from the decompensated category. Creatinine was done for 94(71.2%) and was elevated in more than half 54(57.4%) of the patients, and of those with elevated creatinine, 38(70.4%) were in decompensated stage. Liver function tests were done only in less than one-third (31.8%) of participants and renal function tests were done in only 71.2% participants due to difficulty in obtaining blood sample and severity of shock while presentation, shorter duration of stay in the hospital. The time gap for the initiation of fluid after the patient diagnosed with shock was determined and the mean duration was 24±10minutes. This delay is significant for a shock patient which can contribute to morbidity and mortality, and it may be due to difficult of accessing intravenous line, patient overload, or delay in detecting shock. The frequency of re-evaluation particularly vital signs was determined, and the median was 1hr. Patients were followed after admission and the median length of hospital stay was 7 days. Furthermore, this study showed statistically significant association between presence of underlying illness or chronic medical illness and mortality rate(P<0.001). In conclusion, the prevalence of shock was comparable to other study, but the mortality rate was high where nearly half of the patients had died. Length of illness, type of shock, stage of shock, requirement of mechanical ventilation and duration of hospital stay were significantly associated with bad outcome of shock.

## Declarations

## Acknowledgments

The authors would like to acknowledge medical residents and internship team at Ayder Specialized Hospital, Department of Pediatrics and Child Health for their support in data collection during the study.

## Author Contributions

**Conceptualization:** Mebrahtu G. Kidanu, Alemseged Birhane, Marta Yemane

**Data curation:** Mebrahtu G. Kidanu, Engdaeshet Tazebe

**Formal analysis:** Mebrahtu G. Kidanu, Engdaeshet Tazebe

**Investigation:** Mebrahtu G. Kidanu, Engdaeshet Tazebe

**Methodology:** Mebrahtu G. Kidanu, Engdaeshet Tazebe

**Project administration:** Mebrahtu G. Kidanu

**Software:** Mebrahtu G. Kidanu, Engdaeshet Tazebe

**Supervision:** Alemseged Birhane, Marta Yemane

**Validation:** Mebrahtu G. Kidanu, Engdaeshet Tazebe

**Visualization:** Mebrahtu G. Kidanu, Engdaeshet Tazebe

**Writing original draft:** Mebrahtu G. Kidanu, Mebrahtu G. Tedla

**Writing, review & editing:** Mebrahtu G. Tedla, Mebrahtu G. Kidanu, Mebrihit M. Kahsay, Engdaeshet Tazebe, Alemseged Birhane, Marta Yemane

## Funding information

None

## Data Availability statement

All data related to this study are contained in this paper.

## Competing interests

None

## Notes

### Competing Interest Statement

The authors have declared no competing interest.

### Funding Statement

This study did not receive any funding

### Author Declarations

Institutional Review Board of Mekelle University, College of Health Science gave the ethical approval for this work.

